# Diminished sex hormone levels influence the risk of skewed X chromosome inactivation

**DOI:** 10.64898/2026.04.20.26351303

**Authors:** Amy L. Roberts, Marc F. Östendahl, Aparajita Sahoo, Joseph Pickles, Christopher Franklin-Cheung, Samuel Wadge, Nawal A. Mohamoud, Alessandro Morea, Ariella Amar, David L. Morris, Timothy J. Vyse, Claire J. Steves, Kerrin S. Small

## Abstract

**Background:** X chromosome inactivation (XCI) is the mechanism which randomly silences one X chromosome to equalise gene expression between 46, XX females and 46, XY males. Though XCI is expected to result in a random pattern of mosaicism across tissues, some females display a significantly unbalanced ratio in immune cells, termed XCI-skew, in which ≥75% of cells have the same X inactivated. XCI-skew is associated with adverse health outcomes and its prevalence increases with age – particularly after midlife - yet the specific risk factors have yet to be identified. The menopausal transition, which is driven by profound shifts in sex hormone levels, has significant impact on chronic disease risk yet the molecular and cellular effects are incompletely understood. We hypothesised that the menopausal transition may impact XCI-skew.

**Methods:** Using XCI data measured in blood-derived DNA from 1,395 females from the TwinsUK population cohort, along with questionnaires, genetic data, and sex hormone measures, we carried out a cross-sectional study to assess the impact of the menopausal transition and sex hormones on XCI-skew.

**Results:** We demonstrate that early menopause (<45yrs) is associated with increased risk of XCI-skew. In subset analyses across those who had a surgically induced or natural menopause, we find the association restricted to those who underwent a surgical menopause. We next identify a low polygenic score (PGS) for testosterone levels is significantly associated with XCI-skew, which we replicate in an independent dataset (n=149), while a PGS for age at natural menopause is not associated. Finally, using longitudinal measures across two time points spanning ~18 years we show XCI-skew is a stable cellular phenotype that typically increases over time.

**Discussion:** These data represent the first environmental and genetic risk factors of XCI-skew, both of which implicate endogenous sex hormone levels, particularly testosterone. We propose XCI-skew may have clinical relevance in postmenopausal females.

## Background

The X chromosome is unique in mammalian genetics due to X chromosome inactivation (XCI), which transcriptionally silences all but one X chromosome in each cell to equalise the gene dosage between XX and XY cells ^1^. In humans, the choice of which X to silence in each cell is random and occurs during development, leading to an expected 1:1 ratio of cells with silenced maternal and paternal X chromosomes. The inactive X, termed the Xi, is then clonally inherited by all daughter cells. However, a great deal of variation is observed in immune cells, with some females displaying a “skewed” pattern of XCI (XCI-skew) in which ≥75% of cells have the same Xi ^2^. The proportion of females displaying XCI-skew increases with age and the shifts in XCI ratios across time is thought to reflect long-term changes to the underlying haematopoietic stem and progenitor cells (HSPCs) of the bone marrow ^3,4^.

We have previously shown that XCI-skew correlates with adverse health outcomes and is predictive of future cancer diagnosis ^5^. Intriguingly, XCI-skew is not associated with telomere shortening and epigenetic ageing, suggesting it is an independent cellular marker with clinical potential ^5^. While heritability analyses have shown XCI-skew is influenced by both environmental and inherited genetic factors ^6^, specific risk factors – aside from chronological age - have yet to be identified.

The menopausal transition, which is driven by profound shifts in sex hormone levels and is accompanied by broad physiological changes, has significant impact on chronic disease risk ^7,8^, infectious disease susceptibility, and cellular function^9^. While the average age at menopause is around 50yrs, there is great deal of variability and age at menopause affects disease risk. For example, females who undergo an early menopause (clinically defined as <45yrs) have an augmented risk of cardiovascular disease ^10^. Age at natural menopause (e.g. not surgically induced) has a strong heritable component and genetic variants involved in DNA damage response processes have been identified through GWAS, implicating accelerated biological ageing in the genetic determinants of earlier age at natural menopause ^11^.

In addition, there is growing evidence that endogenous sex hormone levels can impact risk of disease. In females, lower oestrogen exposure is associated with increased risk of dementia ^12^, and lower testosterone is associated with poorer cognition ^13^. Bilateral oophorectomy – the removal of both ovaries – in premenopausal females leads to a sudden drop in oestrogen levels thus inducing the menopause, often with more severe symptoms. It has been shown that while all females with premature menopause (<40yrs) have increased risk of cardiovascular disease, the risk is greatest in those with premature surgical menopause compared to those with a premature natural menopause ^14^. Further, hysterectomy and low testosterone levels have been implicated in risk of frailty ^15^ and mendelian randomisation studies provide evidence of causal links between endogenous sex hormones and cancer risk ^16^.

While the mechanisms linking sex hormones to disease risk are incompletely understood, immune cells may play a key role given sex hormone receptors are expressed across diverse immune cells ^17^. In particular, sex hormone signalling has profound effects on haematopoietic stem and progenitor cells (HSPCs) resulting in shifts in the milieu of mature immune cells produced, as shown by oestrogen-driven changes during pregnancy ^18^ and gender-affirming testosterone therapy in 46, XX transgender men ^19^. A better understanding of immune cell changes driven by sex hormones and the menopausal transition is needed to better understand female health and address the sex bias of age-related morbidity ^20,21^.

Despite many studies exploring XCI across the midlife, the impact of the menopausal transition has yet to be explored. Here we assess the role of menopause and sex hormones on XCI and identify the first genetic and environmental risk factors for XCI-skew. We propose that XCI-skew captures sex hormone-driven changes to HSPCs and holds potential to aid our understanding of sex biases in age-related disease risk.

## Methods

### The TwinsUK XCI cohort

The generation of XCI data using the human androgen receptor assay (HUMARA) and archival blood-derived DNA in the TwinsUK cohort has been previously described ^5^. All samples and information were collected with written and signed informed consent, including consent to publish within the TwinsUK study. TwinsUK has received ethical approval associated with TwinsUK Biobank (19/NW/0187), TwinsUK (EC04/015) or Healthy Ageing Twin Study (HATS) (07 /H0802/84) studies from NHS Research Ethics Service Committees London – Westminster.

Based on the population distribution, a binary variable was created where ≥75% of the cells with the same Xi was defined as XCI-skew (1) and <75% as random XCI (0). Of the 1,575 individuals in the initial study, 79 were excluded due to lack of available genotype data and a further 101 were excluded due to history of cancer prior to sampling (**Supplementary Fig. 1**). This resulted in 1,395 individuals, 98% had self-reported ancestry of white European, with a median age of 60 (IQR 51-67).

Of these, we generated new XCI data in 157 samples assayed at a second time point 14-21 years after the baseline timepoint. Briefly, the HUMARA method was used exactly as described previously using 625ng of genomic DNA across triplicate samples and processed on an ABI 3730xl using the GeneScan 500 LIZ size standard ^5,22^

### Age at menopause and related phenotypic data

Age at menopause was derived from the modal answer from longitudinal self-reported questionnaires. Individuals were defined as post-menopausal if their age at time of sampling was greater than their age at menopause, resulting in 1,020 post-menopausal females, with a median age at menopause of 50 (IQR 45-52; **Supplementary Fig. 2A**). Post-menopausal females were categorised as having experienced an early menopause if their modal age at menopause was <45 and they had never reported an age at menopause ≥45 (n=168; 16.5%). 74 post-menopausal females were excluded due to inconsistent age at menopause reporting, with the remaining 778 experiencing an expected age at menopause (≥45; **Supplementary Fig. 2A**). Self-reported history of hysterectomy and age at hysterectomy data were also used, and surgically induced menopause was defined as 1 year or less between age at menopause and age at hysterectomy, likely reflecting a bilateral oophorectomy (n=196; 19.2%; **Supplementary Fig. 2C**). As expected, those with an induced menopause had a lower age at menopause (median = 42) compared to the group reporting a natural menopause (median age at menopause = 51; **Supplementary Figure 2D**). Further, a subset of 657 participants had self-reported hormone replacement therapy (HRT) use, with 290 and 367 reporting having taken and never having taken HRT respectively, at time of sampling.

### The SLE XCI cohort

The generation of XCI data using the HUMARA assay and archival blood-derived DNA in a cohort of systemic lupus erythematosus (SLE) patients has been previously described ^22^. Of the 181 individuals in the original study, 149 had genome-wide genotype data. All volunteers met the 1997 American College of Rheumatology criteria for SLE ^23^

### Polygenic scores for sex hormones

Summary files for polygenic scores (PGS) for high estradiol [>212 pmol/L] (PGS001182)^24^, testosterone levels in females (PGS001914)^25^ and sex hormone binding globulin (SHBG; PGS001977)^25^ were downloaded from the Polygenic Score Catalogue. Further, publicly available GWAS summary statistics on age at natural menopause were downloaded ^11^. Plink2 was used to calculate the PGS across TwinsUK autosome-wide genotype data (MAF >1%; hg37) using the King’s College London CREATE system ^26^. Given XCI-skew might invalidate the additive assumptions of the PGS model for X-linked SNPs, only autosomal SNPs were included. The PGS for testosterone levels in females (PGS001914) and high estradiol (PGS001182) were also calculated across the SLE autosome-wide genotype data (hg38) for replication. The polygenic score was harmonised to ensure the same variants were captured in both the TwinsUK and SLE genotype datasets. PGSs were normalised and quartiles defined, which were used in all regression models analyses.

### Sex hormone measurements in plasma

Plasma levels of oestradiol, SHBG and testosterone were measured by commercial ElectroChemiLuminescent immunoassays on a Modular Analytics E170 analyser (Roche Diagnostics GmbH, Mannheim, Germany) using the prescribed assay calibrators and performed according to the manufacturer’s protocol. The specific assays used were Estradiol II (03000079; CalSet II 03064921), SHBG (03052001: CalSet 03052028) and Testosterone II (05200067; CalSet II 05202230), as previously described^27^.

### Statistical analysis

Mixed effects regression models were fitted, with family structure and zygosity controlled for as random effects, and age at time of sampling as a fixed covariate. Results from logistic regression models were quantified with odds ratios and 95% CIs. Results from linear regression models were quantified with betas. A paired t-test was used for the discordant twin analyses. All analysis were carried out using R version 4.4.1.

## Results

### Surgically induced early menopause is a risk factor for XCI-skew

Based on the population distribution in TwinsUK, we previously defined XCI-skew as ≥75% of cells having the same Xi ^5^, which is also in line with other published studies ^28^ Given an early menopause (age at menopause <45) is associated with increased disease risk, we explored whether this was a risk factor for XCI-skew. Using a mixed effects logistic regression model controlling for age and relatedness, early menopause (n=168) is associated with increased risk of XCI-skew in post-menopausal females (p=8.3×10^−3^, OR=1.77 (1.16-2.70; **Fig. 1**) compared to those with an expected age at menopause (n=778). Controlling for HRT use in a secondary analysis in the subset with self-reported data (n=657) did not moderate the association with early menopause (p=9.0×10^−3^) and HRT use itself was not associated with XCI-skew (p=0.35).

**Figure 1:**
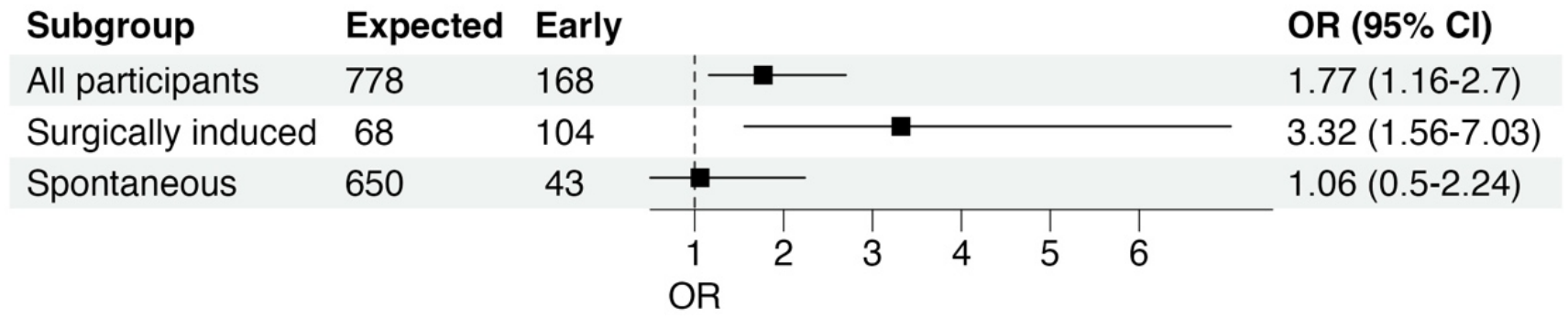
XCI-skew and early menopause. A forest plot showing the odds ratios (OR) and 95% confidence (CI) intervals from logistic regression models for the effect of early menopause (<45yrs) on XCI-skew across: A) all participants (n=946); B) surgically induced menopause (n=172); C) spontaneous menopause (n=693). The line of no effect (OR=1) is shown.

The menopausal transition is accompanied by changes to sex hormone levels as well as broader physiological changes, both of which may impact disease risk. The removal of both ovaries in pre-menopausal females causes a sudden drop in oestrogen, which induces the menopause, and a ~50% decrease in overall testosterone production. Therefore, we next tested whether changes to sex hormones explain the association between early menopause and XCI-skew by stratifying the cohort into those who had a surgically induced menopause (n=196) and those who had never had a hysterectomy (n=731; **Supplementary Fig. 2C**). Within the induced group, early menopause was significantly associated with increased risk of XCI-skew (p=1.8×10^−3^; OR=3.32 (1.56-7.03; **Fig. 1**)) compared to expected age at menopause. However, within the group who never had a hysterectomy, early menopause was not associated with XCI-skew compared to expected age at menopause (p=0.89). Together, these results suggest abrupt disruption to endogenous sex hormone levels, rather than age at natural menopause, impact risk of XCI-skew.

### The genetic determinants of age at natural menopause are not associated with XCI-skew

We next investigated whether earlier age at natural menopause (ANM) was a risk factor for XCI-skew through utilising a polygenic score (PGS; ^11^. The PGS for ANM (n_variants_ = 221) performed as expected in our data and associated significantly with age at menopause in the group who reported never having a hysterectomy (p=1.13×10^−11^; beta=1.41), but not in the group with surgically induced menopause (p=0.83; beta= −0.08, **Supplementary Fig. 3**). However, the ANM PGS was not associated with XCI-skew (n=1,395; p=0.17; **Supplementary Fig. 4A**), supporting our earlier results that the effects of early menopausal transition on XCI-skew were likely driven by sex hormone levels and not age at natural menopause.

### The genetic determinants of sex hormone levels confer risk for XCI-skew

We next wished to further test our hypothesis that sex hormone levels impacted XCI-skew. While cross-sectional measures of plasma levels of testosterone, estradiol, and sex hormone binding globulin (SHBG) were available in the TwinsUK cohort (n=3,265 females; average age = 52), there was poor overlap between these data and the XCI samples. Therefore, we instead used these plasma measurements to validate PGSs for testosterone, estradiol, and SHBG. As expected, the testosterone PGS (n_variants_ = 18,042) was associated with total testosterone (beta=0.13; p<2×10^−16^; **Supp Fig. 5A**), the SHBG PGS (n_variants_ = 50,975) was associated with SHBG levels (beta=0.28; p<2×10^−16^; **Supp Fig. 5B**), and to a lesser extent, the high estradiol PGS (n_variants_ =282) was associated with estradiol measured in plasma (beta= 0.051; p=4.6×10^−4^; **Supp Fig. 5C**).

Using a logistic regression model to test for a linear association of increasing PGS quartiles, we found increasing testosterone PGS is protective against XCI-skew (p=0.023; OR=0.86 (0.76-0.98); **Supplementary Fig. 4B**). Those in the lowest PGS quartile for testosterone levels had a 65% increased risk of XCI-skew compared to the highest quartile (OR=1.65 (1.02-2.67); **Supplementary Fig. 6**). There were no significant associations with the SHBG PGS (p=0.78; **Supplementary Fig. 4C**) or the high estradiol PGS (p=0.43; **Supplementary Fig. 4D**). However, we noted a non-linear trend with the estradiol PGS, where those in both the lowest and highest quartiles have higher levels of XCI-skew compared to the middle 50% (**Supplementary Fig. 4D)**. We assessed this statistically by including the PGS quartiles as categorical variables and found both those in the lowest quartile (p=2.1×10^−3^; OR=1.74 (1.22-2.48)) and highest quartile (p=8.53×10^−5^; OR=2.02 (1.42-2.88)) had increased risk of XCI-skew, compared to the middle quartiles (**Supplementary Fig. 6**).

Given menopausal HRT use impacts endogenous sex hormone levels, including increased estradiol and reduction of testosterone levels (refs), we next carried out subset analyses in post-menopausal females who had no history of HRT use at time of sampling (n=367). Once more, increasing testosterone PGS was protective against XCI-skew (p=4.5×10^−3^; **Fig. 2A**) with those in the lowest PGS quartile having a 2.6-fold increased risk of XCI-skew compared to those in the highest quartile (p=2.9×10^−3^; OR=2.61 (1.38-4.95)). For the estradiol PGS, a non-linear trend was still observed, and both those in the lowest quartile (p=8.8×10^−3^; OR=2.21 (1.22-3.99)) and highest quartile (p=0.014; OR=2.06 (1.16-3.65)) had over a two-fold increased risk of XCI-skew compared to the middle 50%.

**Figure 2:**
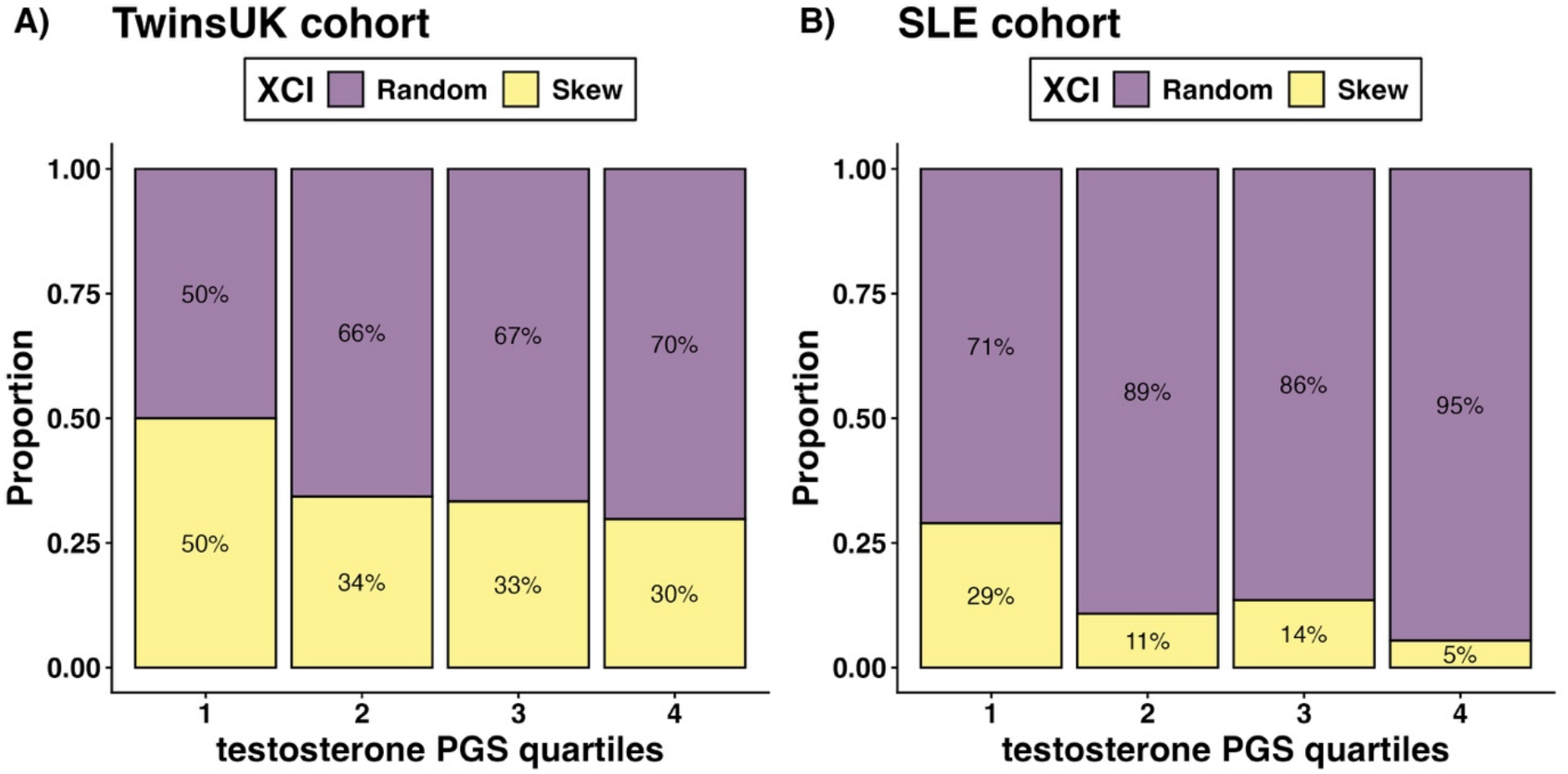
XCI-skew and a testosterone polygenic score. The proportions of individuals (y-axis) with XCI-skew across increasing testosterone polygenic score (PGS) quartiles (x-axis) in A) TwinsUK participants without history of menopausal hormone replacement therapy (HRT) use (n=367) and B) SLE cohort (n=149).

### Replication of the association between testosterone PGS and XCI-skew in a SLE cohort

We have previously shown that XCI-skewing is reduced in systemic lupus erythematosus (SLE) patients and that this was not explained by genetic risk to SLE ^22^. We next used the SLE-XCI cohort (N = 149) for replication. We were able to replicate the association that increasing testosterone PGS is protective against XCI-skew (p=6.2×10^−3^; OR=0.93 (0.89-0.98); **Fig. 2B**), whereas the estradiol PGS did not replicate (p=0.28), even when modelled non-linearly. Of the 22 SLE patients with XCI-skew, 11 are within the lowest quartile for testosterone PGS. Due to the reduced number of SLE patients passing the threshold for XCI-skew ^22^, we also tested the testosterone PGS against XCI-skewing as a continuous variable. Once more, increasing testosterone PGS is protective against increased XCI-skewing (p=0.007; beta = −0.019). Together these results support our hypothesis that endogenous sex hormone levels impact XCI-skew risk, and lower testosterone PGS represents the first genetic risk factor for XCI-skew.

### Surgically induced menopause results in lower testosterone levels

Following the findings from the PGS analysis, we employed a discordant twin approach to explore which hormones may be driving the effects of surgical induced menopause. Within the sex hormone data available in TwinsUK described above (n=3,265 females; average age = 52), we identified 79 postmenopausal monozygotic twin pairs who were discordant for surgically induced or natural menopause, but who of course are perfectly matched for chronological age and inherited genetic factors. Using a paired t-test, we find testosterone levels were lower in those with an induced menopause (p=8.2×10^−3^), whereas there were no significant differences in estradiol (p=0.11) or SHBG (p=0.24). These results support the PGS associations and implicate lower testosterone levels in risk of XCI-skew.

### XCI-skew is a stable cellular phenotype which increases over time

It has previously been demonstrated that XCI measures are stable over short time frames but change over extended periods ^29–31^. To assess the long-term trajectories in an unselected population cohort, we measured XCI in 157 samples at a second time point 14-21 years after the baseline timepoint (median = 18yrs; median age at time point 1 = 53; median age at time point 2 = 70). We defined the XCI categories of random (50-74% of cells with same Xi), skewed (≥75% of cells with same Xi) and extremely skewed (≥91% of cells with same Xi) at both time points. These thresholds were previously calculated based on the population distribution in the TwinsUK XCI cohort ^5^.

Generally, XCI-skew was either stable or increased, as reflected in the median change of 7.4% across the two time points. Of the 112 individuals who had random XCI at time point 1, 76 (68%) remained stable, 31 (28%) progressed to skewed XCI, and five (4%) progressed to extremely skewed XCI (**Fig. 3**). The most extreme change observed across the whole cohort was a 27% increase in XCI representing a shift from 60% (random XCI) of the cells with the same X inactivated to 88% (skewed XCI). Of the 40 individuals with skewed XCI at time point 1, 27 (68%) remained stable, 9 (22%) progressed to extreme skew, and four (10%) decreased to random XCI. This latter group displayed an average change in XCI of −10%. It is very promising to see that XCI-skew may be a reversable cellular phenotype, which is a key aspect of a biomarker, albeit in a small percentage of females.

**Figure 3:**
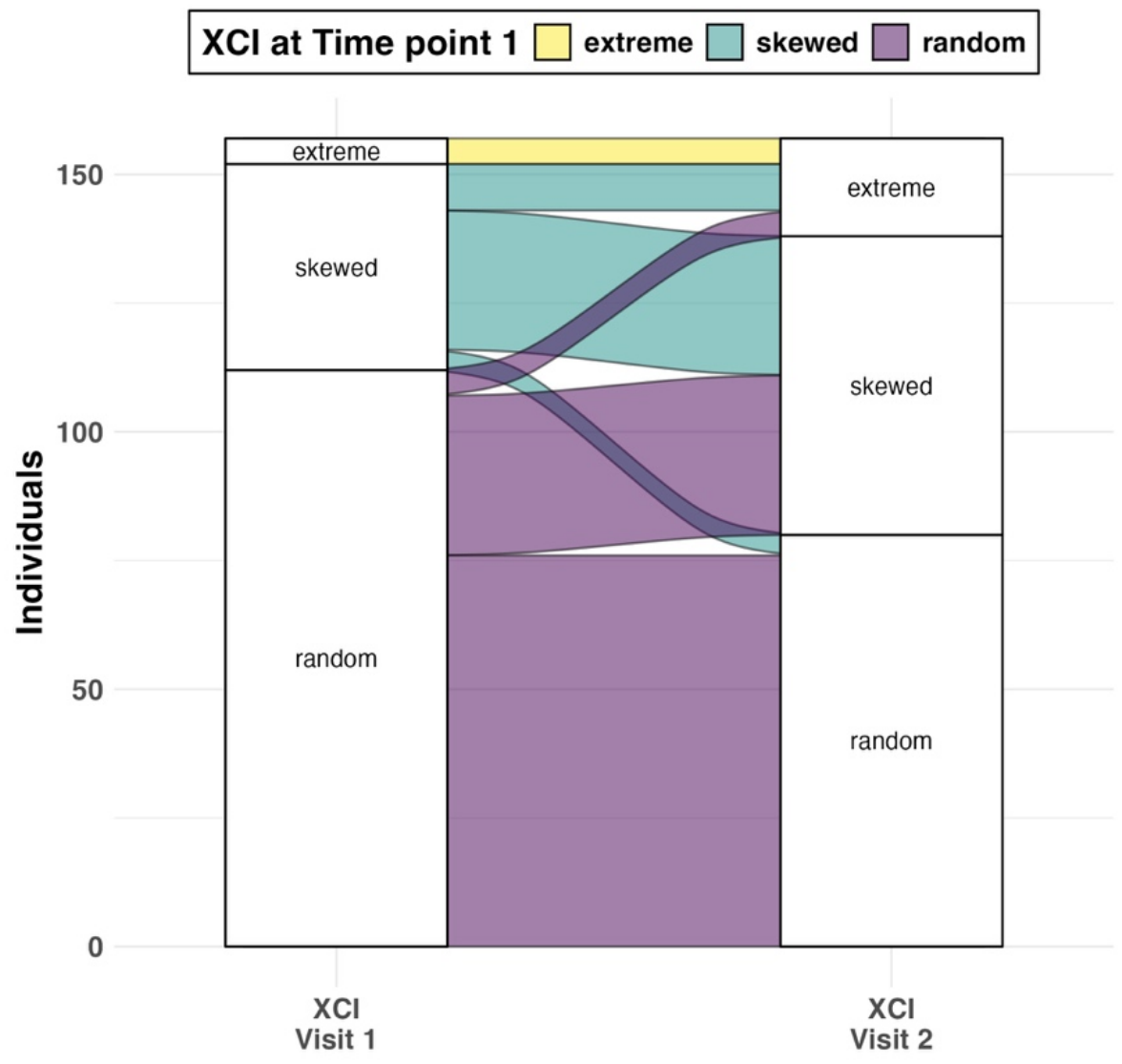
Longitudinal changes in XCI. A Sankey plot showing the longitudinal changes to XCI in 157 individuals across two measures 14-21 years apart (median = 18yrs). Colours indicate XCI at visit 1. Random XCI: 50-74% of cells with same Xi; skewed XCI:≥75% of cells with same Xi; and extremely skewed: ≥91% of cells with same Xi.

To assess whether the menopausal transition impacted how XCI changed longitudinally, we calculated the rate of change of XCI, through dividing the difference in the continuous XCI variable by the difference in years between the two time points. We found no difference in the rate of change of XCI between those who transitioned from pre-to postmenopausal between time points (n=55) compared to who were already postmenopausal at time point 1 (n=95; p=0.52). None of the participants in the premenopausal at time point 1 group had an early menopause and only one had a surgically induced menopause, therefore we were unable to test the effects of these on rate of change of XCI.

## Discussion

XCI-skew is a common cellular phenotype in ageing females associated with adverse health outcomes ^2,4,5^. While heritability analyses suggest both genetic and environmental factors contribute to its development, the specific risk factors for XCI-skew – aside from chronological age - had hitherto been unidentified. Here, using data from 1,544 females across two cohorts, we identify surgically induced early menopause and a low PGS for testosterone levels as environmental and genetic risk factors, respectively, both of which implicate endogenous sex hormone levels, particularly testosterone, in the development of for XCI-skew. Given the profound impact of sex hormones on HSPCs, our work supports earlier hypotheses that XCI-skew captures long-term changes to HSPCs ^3,4^, and we propose XCI-skew may have clinical relevance in postmenopausal females.

While the effects of early menopause, including surgically induced early menopause, on disease risk is appreciated at epidemiological scale ^14^, a better understanding of the molecular and cellular impacts are needed to better predict and prevent disease risk. GWAS studies of age at natural menopause have implicated accelerated biological ageing in the genetic determinants of an earlier age at natural menopause ^11^. Large scale studies in both telomere length shortening and clonal haematopoiesis of indeterminate potential (CHIP) identified associations between earlier age at menopause and these clinically relevant cellular phenotypes ^32,33^. Both studies found these associations were restricted to those with a natural earlier age at menopause, not those with surgically induced early menopause, adding further support to the link between earlier age at natural menopause and accelerated cellular ageing ^32,33^.

Fascinatingly, this is the opposite to what we report here, where surgically induced menopause explained the link between XCI-skew and early menopause, implicating sex hormone levels and not intrinsic cellular ageing as the underlying mechanism. We were able to corroborate this finding by demonstrating a polygenic score for age at natural menopause was not associated with XCI-skew. Together, these findings suggest that XCI-skew is driven by independent mechanisms to CHIP and telomere length thus potentially holding unique clinical utility. This is supported by our earlier finding that telomere length was not associated with XCI-skew ^5^.

We were able to support our mechanistic hypothesis by employing polygenic scores for sex hormone levels, where we find a lower polygenic score for testosterone levels is a risk factor for XCI-skew across two cohorts. These data were supported by a discordant twin model where we find that testosterone levels were significantly lower in individuals who had undergone a surgically induced menopause compared to their monozygotic co-twin who had no history of hysterectomy. This powerful approach, which perfectly controls for chronological age and inherited genetic factors, suggests lower testosterone levels, but not estradiol or SHBG, may be mediating the effects of surgical menopause on XCI-skew. It will be important for future research to determine whether the effect of testosterone on XCI-skew is indeed causative, and whether XCI-skew is a mediator of disease risk, which could support the need for testosterone therapy in postmenopausal females.

Increasing evidence supports the importance of testosterone on female health: low testosterone levels have been linked to lower cognition ^13^ and higher frailty risk ^15^ However, the genetic determinants of testosterone levels reveal a more complex picture. Firstly, a higher polygenic score for testosterone levels was associated with increased metabolic disease in females, whereas it was protective in males, suggesting sex-specific mechanisms ^34^. Although an independent study found no evidence of a causal contribution of sex hormones on metabolic traits^35^. Secondly, a mendelian randomisation study showed evidence for a causal link between total and bioavailable testosterone and reduced risk of ovarian cancers and endometriosis, but increased risk for endometrial cancers ^16^. Ultimately, further research is needed to better understand the cellular and molecular effects of endogenous sex hormones – and the benefit and limitations of hormonal therapies - to uncover the mechanisms of disease risk associated with menopause.

Finally, we demonstrated that XCI is generally stable or increases over a ~18-year window. Although 10% of those with XCI-skew at time point 1 resolved to a random XCI profile by time point 2, it was more common for XCI-skew to progress to extreme skewing (22%), which is in line with earlier research from Danish population cohorts ^31^. We also note that two of the four individuals who resolved from XCI-skew to random XCI – a monozygotic pair – were very close to the lower threshold of XCI-skew at time point 1 (76%), and all 5 individuals with extreme skew at time point 1 still displayed extreme skew at time point 2. The potential of reducing XCI-skew enhances its potential as a cellular biomarker of immune dysfunction and will enable the beneficial environmental factors to be identified. It will be important to replicate this promising finding in a larger cohort and to establish whether those with extreme XCI-skew (>91% of cells with the same Xi) can also be resolved. Further, while our data suggest the menopausal transition did not impact the rate of change of XCI, we were unable to assess the effects of early age at menopause or surgical menopause. Once more, larger longitudinal cohorts will be needed to establish the XCI trajectories dependent on menopausal history.

Our study does have some limitations. Firstly, many of the datasets are derived from self-reported data. Our menopausal status was based on self-reported questionnaire data and not the stages of reproductive aging workshop (STRAW) criteria ^36^. However, we took a robust approach to define age at menopause by using the modal answer across multiple longitudinal questionnaires in TwinsUK. Likewise, when defining the early menopause group, we took a stringent approach and excluded anyone who had ever reported an age at menopause of 45 or more, even if their modal answer fell below 45. Secondly, our definition of surgically induced menopause was defined as 1 year or less between age at menopause and age at hysterectomy. While this definition will capture those who underwent a bilateral oophorectomy, it may also include those who underwent a hysterectomy without the removal of ovaries, which results in the end of menstrual bleeding without inducing the stark reduction in sex hormones. This is likely to have underestimated the true effect of the impact of surgical menopause on XCI-skew. Thirdly, we did not have date-matched measures of sex hormone levels to integrate with the XCI data directly. However, we were able to use these data to demonstrate the effectiveness of the polygenic scores in the wider cohort, which capture lifetime exposure to endogenous sex hormone levels. And finally, while our longitudinal samples were unselected for any phenotypic trait, they were selected on the availability of DNA samples ~18 years apart which could introduce survivor biases into our study. Larger longitudinal studies which avoid such biases are needed.

In summary, our study identifies the first environmental and genetic risk factors for XCI-skew and establishes endogenous sex hormone levels as drivers of this important cellular phenotype. We propose XCI-skew captures long-term changes to HSPCs across the life course, partly driven by sex hormone levels. Our work has important implications for the sex differences in immunity and disease risk, which could help reduce health inequalities across sexes and gender identities.

## Data Availability

All TwinsUK data in the manuscript have been deposited to the TwinsUK BioResource data management team and are available by application to the Twin Research Executive Access committee (TREC) at King’s College London. The TwinsUK BioResource is managed by TREC, which provides governance of access to TwinsUK data and samples. TwinsUK data users are bound by data sharing agreement set out in the data access application form (https://twinsuk.ac.uk/researchers/access-data-and-samples/request-access/). This includes responsibilities with respect to third party data sharing and maintaining participant privacy. Further responsibilities include a responsibility to acknowledge data sharing.

